# Consistency of Linguistic and Cognitive Processing Measures to Discriminate Children with and without Developmental Language Disorder (DLD): Comparing Likelihood Ratios (LHs) and Elastic Net Regression Computational Models

**DOI:** 10.64898/2026.03.09.26347082

**Authors:** Susmi Sharma, Richard M. Golden, James W. Montgomery, Ronald B. Gillam, Julia L. Evans

## Abstract

Because both monothetic and polythetic diagnostic classification approaches focus on the presence of individual symptom(s) to identify individuals in a clinical population, they may be diagnostically sensitive clinical markers of multidimensional disorders such as developmental language disorder (DLD). DLD researchers have also used likelihood ratios (LHs) to identify possible diagnostic clinical markers of DLD, however the diagnostic sensitivity of LHs varies markedly across studies. A recent multidimensional computational elastic-net regression examined a total of 71 measures of spoken language and cognitive processing from a cohort of 223 children ages 7;0 to 11;0 with and without DLD (DLD = 110; typically developing (TD) controls = 113). All 200 iterations of the model had high discriminative power (87% – 88%) in positively identifying and distinguishing the DLD participants across all thresholds. Notably, the models identified a sparse DLD-specific deficit profile which only included nine of the 71 measures. In this study, we ask if the individual LHs for each of these nine measures are equally sensitive in identifying and discriminating the children with DLD from TD controls or if diagnostic markers of multidimensional disorders such as DLD can only be identified based on computational modeling approaches. The LHs for each of the nine measures were in the moderately high ranged (3.25 - 10). However, at the the highest LH cut points for each measure, there was little to no overlap in the children each measure identified as having DLD. Follow up analysis revealed that the elastic net model-derived predictive scores for each participant were significantly correlated with the participants’ language ability. The model also identified a subgroup of TD participants as having the same DLD-deficit profile as the DLD participants. This subgroup were younger, predominantly male participants whose standardized language assessment scores were lower as compared to the larger TD cohort. Taken together, the results from this study show that, because multidimensional modeling approaches such as elastic net regression leverage the variability in the deficit profiles across individual members of a diagnostic group and the unique contributions of each of the behavioral features of the phenotype, they may be an effective tool in deriving diagnostically specific deficit profiles for phenotypically complex, multicausal, multidimensional, neurodevelopmental disorders such as DLD. The results also demonstrate the robustness of the derived DLD-specific deficit profile in identifying individuals with “mild” or subclinical DLD, demonstrating the potential utility of this approach in both clinical and research arenas.

**What this paper adds.:** *What is already known on this subject.:* The identification of diagnostic markers for DLD has been a challenge for both clinicians and researchers across multiple decades. Monothetic classification markers such as non-word repetition, optional infinitive, or syntax dependencies have been explored, as well as polythetic classification approaches where a list of diagnostic symptoms is used together. However, each assumes different criteria and symptoms that should be included as diagnostic markers of DLD.

*What this study adds.:* Our study assessed the feasibility and effectiveness of monothetic vs. polythetic classification approaches for identifying DLD. Since our prior work, which used elastic net logistic regression computational modeling with strong discriminatory power, consistently selected nine key features as the DLD-deficit profile, in this effort, we calculated each of the nine features’ likelihood ratios to examine each measure’s ability to identify children with DLD. The monothetic approach failed to identify a consistent set of children with DLD, and the polythetic classification approach also did not identify participants who were shown to have mild DLD by the elastic net modeling approach. Instead, our analysis showed that a computational modeling approach, such as elastic net regression, that included small but important input from multiple cognitive and linguistic aspects of children, could better capture multifaceted information about the disorder, better account for individual variability, and consistently identify most participants with DLD.

*Clinical implications of this study.:* Elastic net logistic regression identifies a small subset of important features for distinguishing DLD and can assign a probability of DLD presence for each participant. Instead of the polythetic and monothetic approaches commonly used in the field, our study shows that integrating advanced computational modeling, such as elastic net regression, with clinician judgment can better refine assessment processes and address prior and ongoing inconsistencies in the DLD literature and diagnostic practices.

## Introduction

The term Developmental Language Disorder (DLD) refers to a group of children who fail to master spoken and written language comprehension and production despite normal hearing acuity and the absence of any medical condition or syndrome known to cause language disorders in children (Leonard, 2014). If undiagnosed and/or untreated, persists into adulthood with significant consequences throughout the individual’s life span (Botting et al., 2016; Conti-Ramsden et al., 2013; Gaëlle Hubert-Dibon et al., 2016). The objective identification of DLD and the differentiation of individuals with DLD from typically developing (TD) controls has been a longstanding challenge for clinicians and researchers. Although there is consensus that the disorder is characterized by impaired language comprehension and production and general agreement in the use of exclusionary diagnostic criteria (e.g., absence of hearing impairment, social-emotional deficits, and motor impairment), there is a lack of consensus regarding which inclusionary markers best identify these individuals.

Polythetic diagnostic approaches classify a disorder from a list of symptoms, where a child’s diagnosis is based on the presence of a predefined subset of those symptoms. The EpiSLI diagnostic system proposed by Tomblin and colleagues is an example of this type of approach (Tomblin et al., 1996) where they classified children based on five measures from three language domains (vocabulary, grammar, and narration) and two modalities (comprehension and production). Children having 2:5 scores at or below -1.25 standard deviations were classified as having DLD. Because a polythetic classification approach relies on the presence/absence of a list of symptoms, however, it can artificially create the appearance of heterogeneous deficit profile for a clinical population. For example, if 5:9 symptoms are required for a child be classified with a given disorder, there will be a total of 150 different possible combinations of the symptoms and any two children who meet the 5:9 criteria may end up having only one symptom in common.

Alternatively, the monothetic classification approach defines a disorder based on a single diagnostic symptom. Rice and colleagues’ proposal that omission of optional infinitive is the defining marker of DLD (Rice et al., 2023) or van der Lely and colleagues’ proposal that impaired syntactic dependencies involving “movement” as in filler-gap dependencies are diagnostic markers of DLD (Marinis and van der Lely, 2007) are examples of this approach. A disadvantage of a monothetic diagnostic classification approach is that diagnostic features are linked to different theoretical account of a disorders. For example, while Rice and van der Lely argue that the defining diagnostic feature of DLD is linked to aspects of the linguistic system, other researchers have argued that the defining diagnostic feature of DLD may lie in the nonlinguistic cognitive domains. The focus on phonological working memory as a diagnostic marker of DLD based on nonword repetition tasks (NRT) is such as example. For example, Dollaghan & Campbell (1998), Ellis Weismer et al. (2000), Girbau & Schwartz (2007), and Ahufinger et al. (2021) NRT using positive likelihood ratios to identify cut-points in children’s performance to differentiate children with DLD from children with normal language abilities. The results of these studies are inconsistent, however, both with respect to the degree to which cut off points in the children’s performance differentiated children with DLD from typical language controls *and* in the actual positive likelihood ratio values reported across studies.

Positive likelihood ratios (also referred to as LH+) are a measure of the percentage possibility of an individual being a part of a group given their score on a task or test (Tucker and Lewis, 1973). The larger the LH value, the greater the likelihood the individual whose performance falls in a given range is a member of the diagnostic group. The LHs values for these NRT studies ranged from small to moderate (LH = 2.78 at or below 70% accuracy, Ellis Weismer et al., 2000), moderate (LH = 6.67 at or below 50% accuracy, Girbau and Schwartz, 2007) to moderate-high (LH = 11 at or below 50% accuracy, Girbau and Schwartz, 2007). Recently, Ahufinger and colleagues asked if NRT could be used to classify Catalan–Spanish and European Portuguese-speaking children with and without DLD (Ahufinger et al., 2021). Their LHs were higher than previously reported for their Catalan-Spanish speaking and European Portuguese speaking children respectively (LH+ = 33; LH+ = 35) at 70% and 75% total phonemes correct cut-points, due in part to differences in the wordlikeness of their stimuli, indicating that even for the same task, LH values can also be sensitive to stimuli factors such as the wordlikeness or syllable length of the nonwords.

Another limitation of both polythetic and monothetic classification approaches is their inability to account for different underlying etiologies, different ages of onset, or severity or the presence of underlying unobservable dimensions of the disorder. The problem of classification is further complicated for these approaches if a “symptom” is qualitative in nature, varies along the continuous normal distribution, and/or has a high degree of overlap in the observable traits of the disorder and unimpaired individuals, which requires the use of an arbitrary cutoff score for a symptom to be considered “present.” Taken together, these challenges raise significant questions regarding the feasibility of these approaches in accurately diagnosing children with DLD. DLD, similar to other neurodevelopmental disorders, such as autism spectrum disorder (ASD), is characterized by a wide range of types and severity of symptoms that may not be expressed by different individuals but may also be expressed by the *same* individual at different stages in development. It may also be a multi-causal disorder with comorbid conditions. The problem in identifying children with DLD is compounded further by the fact that the various language measures and cognitive tasks have been conceptualized and measured as dimensional, when in fact, at the latent level, they may not be dimensional along the entire continuum. As a result, dimensional variation may be present in those children who fall along the normal/upper range of the language continuum, but children who fall at the extreme low end of the continuum may be categorically distinct (Ruscio et al., 2011).

In a recent computational modeling study, we modeled DLD as a multi-dimensional disorder and used a repeated elastic net regression machine learning algorithm approach to ask if there might be a DLD-specific deficit profile could be identified based on an extremely large number of theoretically motivated cognitive and language measures (Sharma et al., in press; Sharma et al., submitted for publication). We examined 71 measures of real-time spoken sentence comprehension, lexical processing, working memory, attentional control, and speed of processing from the 117 children with DLD and 117 typically developing (TD) controls (ages 7 – 11) from the large-scale multi-site Montgomery et al.’s study (Montgomery et al., 2017). Our model distinguished 87%-88% of the children as having DLD across all thresholds. Tracking which features the models used to distinguish the DLD and TD children over the 200 iterations gave us a weighted index of each of the 71 features’ importance (Weighted Feature Important Index (WFII)) in identifying a DLD-specific profile. Surprisingly, our models used a total of fourteen measures. Of these fourteen measures a subset of *nine* language and cognitive processing measures were used by the model to identify DLD. The nine measures that comprised the unique DLD-deficit profile included both linguistic measures of spoken sentence comprehension and several different cognitive measures of verbal working memory, phonological working memory and lexical processing speed.

### Current Study

In this study, we ask if LH+ can also discriminate these children with and without DLD or if computational modeling approaches (e.g., elastic net regression model) are needed to identify the unique disorder-specific features of multidimensional disorders such as DLD. We derived LHs values for each of the nine measures from the DLD-deficit profile and re-examined the discrimination of each of them based on their positive LHs (e.g., ability to identify the DLD participants).

## Method

### Participants

The participants consisted of 234 children (ages 7:0 to 11:11), 117 children with DLD (72 boys and 45 girls), and 117 children with typical language (TD) (83 boys and 34 girls) from the Montgomery et al., (2017) study. In the original Montgomery et al study, the children were classified as DLD-TD based on scores from four standardized assessment measures: the receptive and expressive portions of the Comprehensive Receptive and Expressive Vocabulary Test (CREVT-2; Wallace and Hammill, 2000), and the concepts and following directions subtest and recalling sentences subtest of the Clinical Evaluation of Language Fundamentals (CELF-4; Semel et al., 2003). Children were classified as DLD if their mean composite language z-score on the three lowest of the four measures was at or below -1. Children were classified as typically developing if their average composite z-score was greater than -1.

### Elastic Net Regression Model

For our elastic net regression model, we used all of the experimental measures of spoken sentence comprehension and cognitive processing from the Montgomery study and the Gillam-Evans-Montgomery (GEM) model (Montgomery et al. 2017; Montgomery et al. 2016; Montgomery et al. 2018). These included 71 measures of canonical and noncanonical spoken sentence comprehension, lexical processing, including rapid automatic naming and spoken word recognition, controlled attention, phonological short-term memory, and verbal and nonverbal working memory. We used the “cv.glmnet” function from the glmnet package in R (Friedman et al., 2010; Friedman et al., 2023), which combines elastic net logistic regression with 10-fold cross-validation to find the optimal λ that results in a low-dimensional set of features reliably predicting the presence of DLD. We trained the model a total of 200 times, resulting in 200 fitted models for analysis. For detailed description of the model and the 71 measures please see Sharma et al (2025; in press, under review).

The number of times each of the 71 features was selected across the 200 runs was computed which enabled us to identify which of the 71 features were consistently used to classify DLD and TLC participants across all 200 runs. The model removed 87.4% of the input variables, consistently retaining only nine measures. These included: (1) Total Spoken Sentence Comprehension Accuracy (TotSyntaxComAcc), (2) Noncanonical Sentence Comprehension Accuracy (NonCanonicalAcc), (3) Passive Sentence Comprehension Accuracy (PasAcc), (4) Nonword Repetition Total Phonemes correct (Dollaghan_PPC), (5) Working Memory Trials Correct (WMTrialsCor), (6) Working Memory Span (WmSpan), (7) Digit Span Trials Correct (DigitSpTrialsCor), (8) Digit Span (Digit_Span), and (9) Speed of Rapid Automatic Naming (RANMeanRT).

In the current study, we also used the novel approach to develop a scoring system to quantify the DLD-deficit profile by examining the model-predicted scores against participants’ composite diagnostic scores. For each model iteration, each participant was assigned a predicted probability score. This predicted probability score was the model’s estimate of DLD presence for each participant. We examined the relationship between DLD status based on the participant’s composite diagnostic score and the DLD-specific profile derived from the models.

### Positive Likelihood Ratios (LHs+) to Rule In Developmental Language Disorder

We calculated the LH+ using the presence/absence of DLD to determine the ability of each of the nine measures to diagnose children with DLD (Sackett et al., 1991; Haynes et al., 2006). To determine the LH+ ratio, the true positive rate (proportion of children with DLD with a score at or below x-determined cutoff) was divided by the false positive rate (proportion of TD children with a score at or below x-determined cutoff). We used Haynes et al. (2006) criteria to classify a positive test (i.e., accurately ruling in the disorder), which includes the following: (1) “High” as defined as LH ratio of 20 or higher having a probability of 95% or greater that the disorder is present, (2) “Intermediate High” defined as an LH+ ratio between 1 and 20, and (3) “Indeterminate” defined as an LH close to 1.0. To calculate cutoff scores that maximize the ability to “rule in” DLD, we calculated the number and proportion of children in the DLD and TD groups whose scores were at or within a given feature value (test-positive).

## Results

The means, standard deviations, and Cohen’s *d* for the DLD and TD participants for the nine measures are shown in Table 1. Figures 1 show each participants’ composite language z-scores plotted against their performance for each of the nine measures. Figure 2 shows a hypothetical plot of the separation of the two groups with a high degree of accuracy (e.g., minimal overlap). As can been seen in Figure 1, although the mean separability is visible for the DLD and TD groups. there is also a large degree of overlap for the two groups.

**Figure 1.**
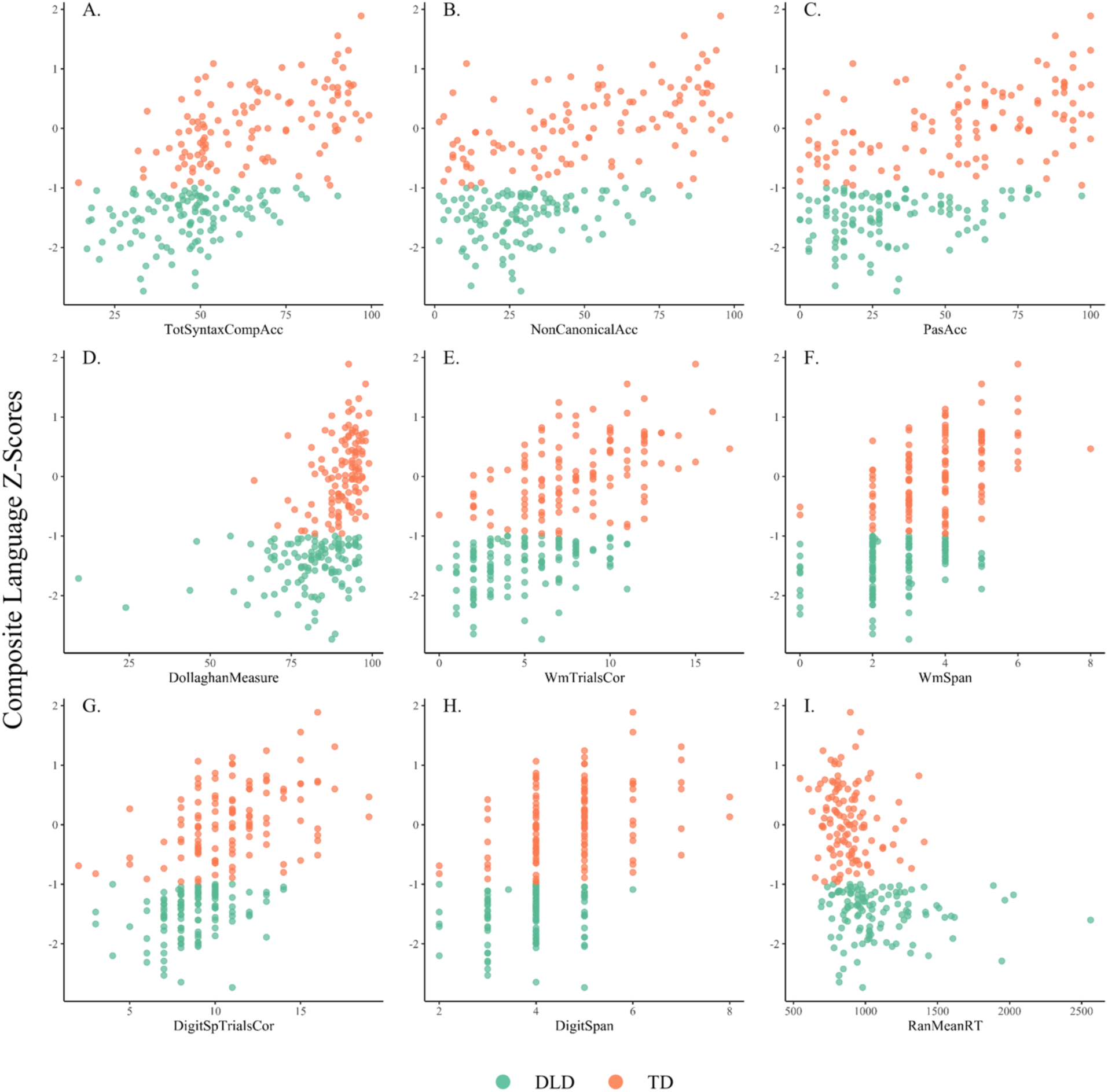
Participants’ Scores in the Nine Measures and their Composite Language z-scores across groups. Figure 1 (A-I) displays the relationship between participants’ scores on each measure and their composite language z-scores. Each point represents a participant in the dataset, with DLD in green and TD in orange. None of the distributions of the 9 measures show a cutoff point that separates participants with DLD from those with typical language. Participants from both groups were presented across the full score range.

**Figure 2.**
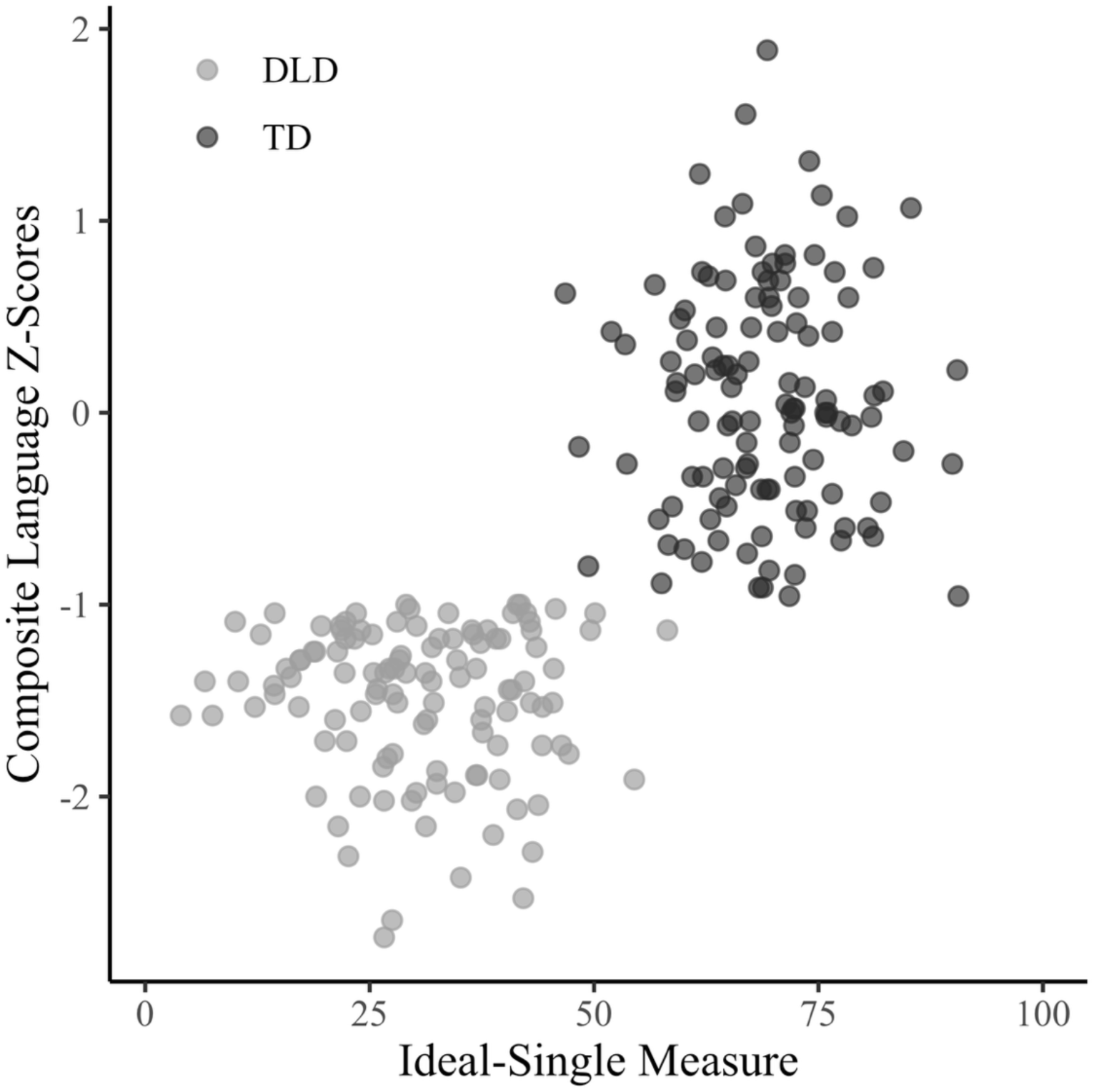
Hypothetical Single Ideal Measure Separation Illustration. In the presence of an ideal, perfect univariate measure able to discriminate participants with DLD from TD, the scatterplot of DLD and TD participants’ score distribution should cleanly separate into two groups, as in the figure.

**Table 1.**
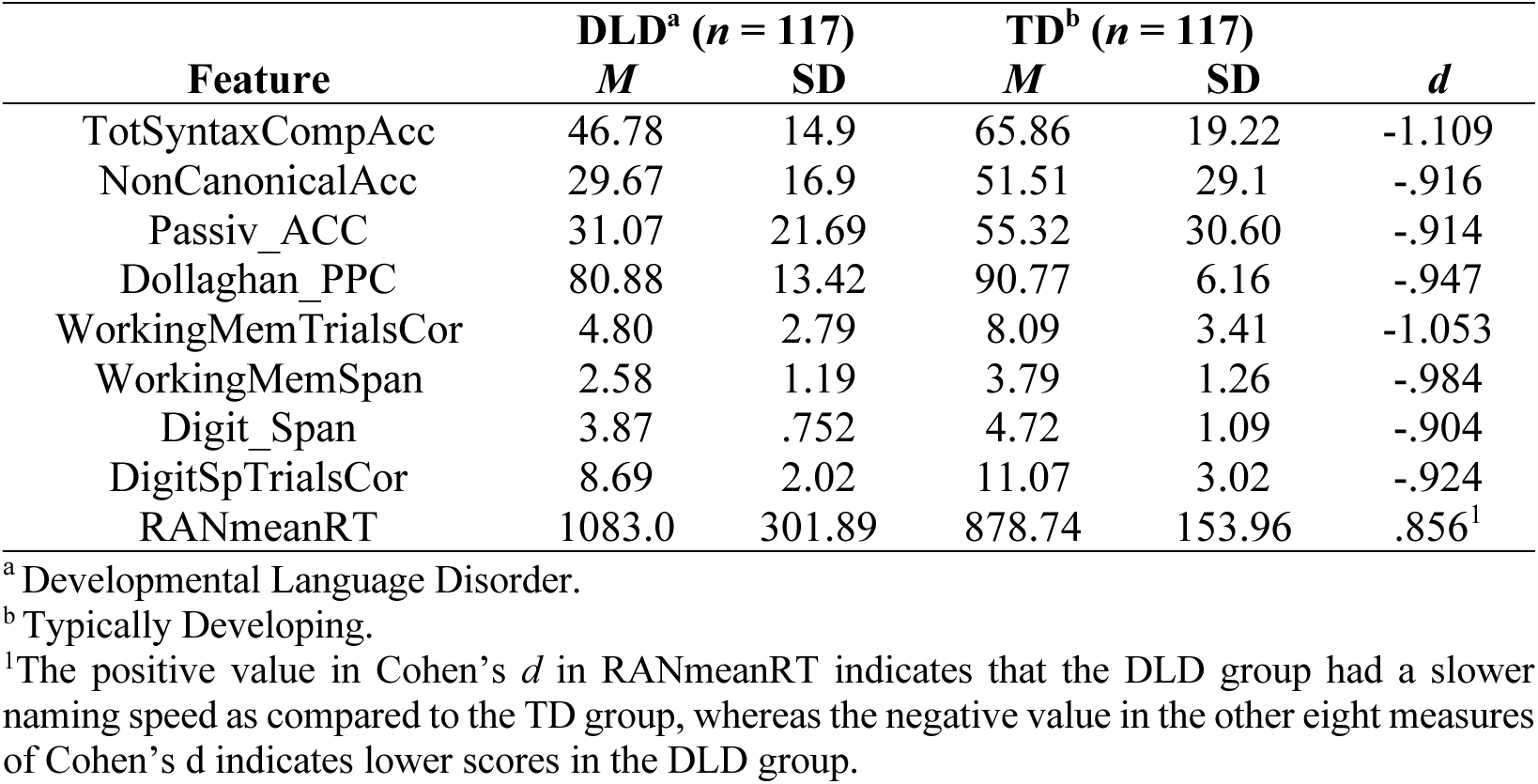
Means, standard deviation, and Cohen’s *d* for the nine measures, shown to be highly important in distinguishing participants with DLD from those with typical language.

As can be seen in Figure 2, an ideal measure would have an obvious cutoff value (e.g., 50 in this case), where the scores for the participants with DLD would all fall below a given cutoff and the scores for the participants with TD would all be above the cutoff. However, as can be seen in Figure 1, there is no single cutoff value for the nine measures for which participants’ distribution of scores aligns with the plot in Figure 2, where the participants with DLD fall on one side and the TD group on the other. This suggests that the nine measures individually may not differentiate the majority of the DLD children from the TD children.

### Rule in DLD based on Positive Likelihood Ratios

The number and proportion of children with a positive test result, sensitivity, specificity, and Positive LHs for the nine measures from the DLD-specific deficit profile are reported in Tables 2 - 10. The LHs were all in the intermediate high range. Digit Span Trials Correct had the highest LH+ (LH ratio of 10) at a cut point of 7 trials correct (DLD = 20; TD = 2); whereas Digit Span had the lowest LH+ (LH ratio of 3.25) at a cut point of a digit span of 3 (DLD = 26; TD = 8).

**Table 2.** Likelihood (LH) ratio analysis for Total Sentence Accuracy for the 117/117 Cohort.

| Tot PSC | <u>DLD<sup>a</sup></u> |  | <u>TD<sup>b</sup></u> |  | Sensitivity | Specificity | LH+ |
| --- | --- | --- | --- | --- | --- | --- | --- |
|  | No. | Prop. | No. | Prop. |  |  |  |
| 0-10% | 0 | 0.000 | 0 | 0.000 | 0.000 | 1.000 | Inf. |
| **11-20%** | **6** | **0.051** | **1** | **0.009** | **0.051** | **0.991** | **6.000\*** |
| 21-30% | 11 | 0.094 | 0 | 0.000 | 0.094 | 1.000 | Inf. |
| 31-40% | 21 | 0.180 | 6 | 0.051 | 0.179 | 0.949 | 3.500 |
| 41-50% | 39 | 0.333 | 28 | 0.239 | 0.333 | 0.761 | 1.393 |
| 51-60% | 21 | 0.180 | 17 | 0.145 | 0.179 | 0.855 | 1.235 |
| 61-70% | 11 | 0.094 | 20 | 0.171 | 0.094 | 0.829 | 0.550 |
| 71-80% | 6 | 0.051 | 10 | 0.086 | 0.051 | 0.915 | 0.600 |
| 81-90% | 2 | 0.017 | 22 | 0.188 | 0.017 | 0.812 | 0.091 |
| 91-100% | 0 | 0.000 | 13 | 0.111 | 0.000 | 0.889 | 0.000 |
| <u>Extreme Cut Points</u> |  |  |  |  |  |  |  |
| **0-35%** | **29** | **0.248** | **5** | **0.043** | **0.248** | **0.957** | **5.800\*** |
| 36-75% | 84 | 0.718 | 73 | 0.624 | 0.718 | 0.376 | 1.151 |
| 76-100% | 4 | 0.034 | 39 | 0.333 | 0.034 | 0.667 | 0.103 |

<sup>a</sup> Developmental Language Disorder.<sup>b</sup> Typically Developing.
\*The cutoffs for the best LH+ ratio for the measure are highlighted in bold. All scores with decimal values were rounded down to the nearest integer and were part of the corresponding range.

**Table 3.** Likelihood (LH) ratio analysis for NonCanonical Sentence Accuracy for the 117/117 Cohort.

| TOT PSC | <u>DLD<sup>a</sup></u> |  | <u>TD<sup>b</sup></u> |  | Sensitivity | Specificity | LH+ |
| --- | --- | --- | --- | --- | --- | --- | --- |
|  | No. | Prop. | No. | Prop. |  |  |  |
| 0-10% | 15 | 0.128 | 15 | 0.128 | 0.128 | 0.872 | 1.000 |
| 11-20% | 20 | 0.171 | 10 | 0.086 | 0.171 | 0.915 | 2.000 |
| **21-30%** | **36** | **0.308** | **5** | **0.043** | **0.308** | **0.957** | **7.200\*** |
| 31-40% | 23 | 0.197 | 15 | 0.128 | 0.197 | 0.872 | 1.533 |
| 41-50% | 9 | 0.077 | 12 | 0.103 | 0.077 | 0.897 | 0.750 |
| 51-60% | 7 | 0.060 | 12 | 0.103 | 0.060 | 0.897 | 0.583 |
| 61-70% | 4 | 0.034 | 11 | 0.094 | 0.034 | 0.906 | 0.364 |
| 71-80% | 2 | 0.017 | 9 | 0.077 | 0.017 | 0.923 | 0.222 |
| 81-90% | 1 | 0.009 | 19 | 0.162 | 0.009 | 0.838 | 0.053 |
| 91-100% | 0 | 0.000 | 9 | 0.077 | 0.000 | 0.923 | 0.000 |
| <u>Extreme Cut Points</u> |  |  |  |  |  |  |  |
| **0 – 30%** | **71** | **0.607** | **30** | **0.256** | **0.607** | **0.744** | **2.367\*** |
| 31-70% | 43 | 0.368 | 50 | 0.427 | 0.368 | 0.573 | 0.860 |
| 71-100 | 3 | 0.026 | 37 | 0.316 | 0.026 | 0.684 | 0.081 |

<sup>a</sup> Developmental Language Disorder.
<sup>b</sup> Typically Developing.
\*The cutoffs for the best LH+ ratio for the measure are highlighted in bold. All scores with decimal values were rounded down to the nearest integer and were part of the corresponding range.

**Table 4.** Likelihood (LH) ratio analysis for Passive Sentence Accuracy for the 117/117 Cohort.

| TOT PSC | <u>DLD<sup>a</sup></u> |  | <u>TD<sup>b</sup></u> |  | Sensitivity | Specificity | LH+* |
| --- | --- | --- | --- | --- | --- | --- | --- |
|  | No. | Prop. | No. | Prop. |  |  |  |
| 0-10% | 17 | 0.145 | 14 | 0.120 | 0.145 | 0.880 | 1.214 |
| 11-20% | 26 | 0.222 | 11 | 0.094 | 0.222 | 0.906 | 2.364 |
| **21-30%** | **29** | **0.248** | **5** | **0.043** | **0.248** | **0.957** | **5.800** |
| 31-40% | 14 | 0.120 | 8 | 0.068 | 0.120 | 0.932 | 1.750 |
| 41-50% | 6 | 0.051 | 6 | 0.051 | 0.051 | 0.949 | 1.000 |
| 51-60% | 11 | 0.094 | 19 | 0.162 | 0.094 | 0.838 | 0.579 |
| 61-70% | 6 | 0.051 | 13 | 0.111 | 0.051 | 0.889 | 0.462 |
| 71-80% | 6 | 0.051 | 9 | 0.077 | 0.051 | 0.923 | 0.667 |
| 81-90% | 1 | 0.009 | 17 | 0.145 | 0.009 | 0.855 | 0.059 |
| 91-100% | 1 | 0.009 | 15 | 0.128 | 0.009 | 0.872 | 0.067 |
| <u>Extreme Cut Points</u> |  |  |  |  |  |  |  |
| **0 – 30%** | **72** | **0.615** | **30** | **0.256** | **0.615** | **0.744** | **2.400** |
| 51-70% | 17 | 0.145 | 32 | 0.274 | 0.145 | 0.727 | 0.531 |
| 71-100 | 8 | 0.068 | 41 | 0.350 | 0.068 | 0.650 | 0.195 |

<sup>a</sup> Developmental Language Disorder.
<sup>b</sup> Typically Developing.
\*The cutoffs for the best LH+ ratio for the measure are highlighted in bold. All scores with decimal values were rounded down to the nearest integer and were part of the corresponding range.

**Table 5.** Likelihood (LH) ratio analysis for Total Phonemes Percent Correct Dollaghan for the 117/117 Cohort.

| Tot PSC | <u>DLD<sup>a</sup></u> |  | <u>TD<sup>b</sup></u> |  | Sensitivity | Specificity | LH+ |
| --- | --- | --- | --- | --- | --- | --- | --- |
|  | No. | Prop. | No. | Prop. |  |  |  |
| 0-10% | 1 | 0.009 | 0 | 0.000 | 0.009 | 1.000 | Inf. |
| 11-20% | 0 | 0.000 | 0 | 0.000 | 0.000 | 1.000 | Inf. |
| 21-30% | 1 | 0.009 | 0 | 0.000 | 0.009 | 1.000 | Inf. |
| 31-40% | 0 | 0.000 | 0 | 0.000 | 0.000 | 1.000 | Inf. |
| 41-50% | 2 | 0.017 | 0 | 0.000 | 0.017 | 1.000 | Inf. |
| 51-60% | 2 | 0.017 | 0 | 0.000 | 0.017 | 1.000 | Inf. |
| **61-70%** | **14** | **0.119** | **2** | **0.017** | **0.1196** | **0.9914** | **7.000** |
| 71-80% | 26 | 0.222 | 6 | 0.051 | 0.2222 | 0.9401 | 4.333 |
| 81-90% | 50 | 0.427 | 40 | 0.342 | 0.427 | 0.658 | 1.250 |
| 91-100% | 21 | 0.180 | 69 | 0.590 | 0.179 | 0.410 | 0.304 |
| <u>Extreme Cut Points</u> |  |  |  |  |  |  |  |
| 0-60% | 6 | 0.051 | 0 | 0.000 | 0.051 | 1.000 | Inf. |
| **61-80** | **40** | **0.342** | **8** | **0.068** | **0.342** | **0.932** | **5.000** |
| 81-100 | 71 | 0.607 | 109 | 0.932 | 0.607 | 0.068 | 0.651 |

<sup>a</sup> Developmental Language Disorder.
<sup>b</sup> Typically Developing.
\*The cutoffs for the best LH+ ratio for the measure are highlighted in bold. All scores with decimal values were rounded down to the nearest integer and were part of the corresponding range.

**Table 6.** Likelihood (LH) ratio analysis for Working Memory Trials Correct for the 117/117 Cohort.

| TOT COR | <u>DLD<sup>a</sup></u> |  | <u>TD<sup>b</sup></u> |  | Sensitivity | Specificity | LH+ |
| --- | --- | --- | --- | --- | --- | --- | --- |
|  | No. | Prop. | No. | Prop. |  |  |  |
| 0 | 1 | 0.009 | 1 | 0.009 | 0.009 | 0.991 | 1.000 |
| 1 | 8 | 0.068 | 0 | 0.000 | 0.068 | 1.000 | Inf. |
| 2 | 23 | 0.197 | 7 | 0.060 | 0.197 | 0.940 | 3.286 |
| **3** | **19** | **0.162** | **4** | **0.034** | **0.162** | **0.966** | **4.750** |
| 4 | 8 | 0.068 | 2 | 0.017 | 0.068 | 0.983 | 4.000 |
| 5 | 13 | 0.111 | 12 | 0.103 | 0.111 | 0.897 | 1.083 |
| 6 | 10 | 0.086 | 14 | 0.120 | 0.085 | 0.880 | 0.714 |
| 7 | 12 | 0.103 | 15 | 0.128 | 0.103 | 0.872 | 0.800 |
| 8 | 11 | 0.094 | 10 | 0.086 | 0.094 | 0.915 | 1.100 |
| 9 | 2 | 0.017 | 10 | 0.086 | 0.017 | 0.915 | 0.200 |
| 10 | 7 | 0.060 | 12 | 0.103 | 0.060 | 0.897 | 0.583 |
| 11 | 3 | 0.026 | 8 | 0.068 | 0.026 | 0.932 | 0.3750 |
| 12 | 0 | 0.000 | 13 | 0.111 | 0.000 | 0.889 | 0.000 |
| 13 | 0 | 0.000 | 3 | 0.026 | 0.000 | 0.974 | 0.000 |
| 14 | 0 | 0.000 | 2 | 0.017 | 0.000 | 0.983 | 0.000 |
| 15 | 0 | 0.000 | 2 | 0.017 | 0.000 | 0.983 | 0.000 |
| 16 | 0 | 0.000 | 1 | 0.009 | 0.000 | 0.991 | 0.000 |
| 17 | 0 | 0.000 | 1 | 0.009 | 0.000 | 0.991 | 0.000 |
| <u>Extreme Cut Points</u> |  |  |  |  |  |  |  |
| **≤4** | **51** | **0.436** | **12** | **0.103** | **0.436** | **0.897** | **4.250** |
| 7-10 | 56 | 0.479 | 63 | 0.539 | 0.479 | 0.462 | 0.889 |
| ≥11 | 10 | 0.086 | 42 | 0.359 | 0.085 | 0.641 | 0.238 |

<sup>a</sup> Developmental Language Disorder.
<sup>b</sup> Typically Developing.
\*The cutoffs for the best LH+ ratio for the measure are highlighted in bold. All scores with decimal values were rounded down to the nearest integer and were part of the corresponding range.

**Table 7.** Likelihood (LH) ratio analysis for Working Memory Span for the 117/117 Cohort.

| Span | <u>DLD<sup>a</sup></u> |  | <u>TD<sup>b</sup></u> |  | Sensitivity | Specificity | LH+ |
| --- | --- | --- | --- | --- | --- | --- | --- |
|  | No. | Prop. | No. | Prop. |  |  |  |
| **0** | **10** | **0.086** | **2** | **0.017** | **0.085** | **0.983** | **5.000** |
| 2 | 52 | 0.444 | 14 | 0.120 | 0.444 | 0.880 | 3.714 |
| 3 | 28 | 0.239 | 31 | 0.265 | 0.239 | 0.735 | 0.903 |
| 4 | 21 | 0.180 | 38 | 0.325 | 0.179 | 0.675 | 0.553 |
| 5 | 6 | 0.051 | 23 | 0.197 | 0.051 | 0.803 | 0.261 |
| 6 | 0 | 0.000 | 8 | 0.068 | 0.000 | 0.932 | 0.000 |
| 7 | 0 | 0.000 | 0 | 0.000 | 0.000 | 1.000 | Inf. |
| 8 | 0 | 0.000 | 1 | 0.009 | 0.000 | 0.991 | 0.000 |
| <u>Extreme Cut Points</u> |  |  |  |  |  |  |  |
| **<2** | **62** | **0.530** | **16** | **0.137** | **0.530** | **0.863** | **3.875** |
| 3,4 | 49 | 0.419 | 69 | 0.590 | 0.419 | 0.410 | 0.710 |
| >5 | 27 | 0.051 | 32 | 0.274 | 0.051 | 0.727 | 0.188 |

<sup>a</sup> Developmental Language Disorder.
<sup>b</sup> Typically Developing.
\*The cutoffs for the best LH+ ratio for the measure are highlighted in bold. All scores with decimal values were rounded down to the nearest integer and were part of the corresponding range.

**Table 8.** Likelihood (LH) ratio analysis for Digit Span Trials Correct for the 117/117 Cohort.

| Span | <u>DLD<sup>a</sup></u> |  | <u>TD<sup>b</sup></u> |  | Sensitivity | Specificity | LH+ |
| --- | --- | --- | --- | --- | --- | --- | --- |
|  | No. | Prop. | No. | Prop. |  |  |  |
| 1 | 0 | 0.000 | 0 | 0.000 | 0.000 | 1.000 | Inf. |
| 2 | 0 | 0.000 | 1 | 0.009 | 0.000 | 0.991 | 0.000 |
| 3 | 2 | 0.017 | 1 | 0.009 | 0.017 | 0.991 | 2.000 |
| 4 | 2 | 0.017 | 0 | 0.000 | 0.017 | 1.000 | Inf. |
| 5 | 1 | 0.009 | 3 | 0.026 | 0.009 | 0.974 | 0.333 |
| 6 | 5 | 0.043 | 1 | 0.009 | 0.043 | 0.991 | 5.000 |
| **7** | **20** | **0.171** | **2** | **0.017** | **0.171** | **0.983** | **10.000** |
| 8 | 27 | 0.231 | 8 | 0.060 | 0.231 | 0.932 | 3.375 |
| 9 | 23 | 0.197 | 21 | 0.180 | 0.197 | 0.821 | 1.095 |
| 10 | 20 | 0.171 | 14 | 0.120 | 0.171 | 0.880 | 1.429 |
| 11 | 7 | 0.060 | 21 | 0.180 | 0.060 | 0.821 | 0.333 |
| 12 | 4 | 0.034 | 13 | 0.111 | 0.034 | 0.889 | 0.308 |
| 13 | 4 | 0.034 | 9 | 0.077 | 0.034 | 0.923 | 0.444 |
| 14 | 2 | 0.017 | 5 | 0.043 | 0.017 | 0.957 | 0.400 |
| 15 | 0 | 0.000 | 7 | 0.060 | 0.000 | 0.940 | 0.000 |
| 16 | 0 | 0.000 | 7 | 0.060 | 0.000 | 0.940 | 0.000 |
| 17 | 0 | 0.000 | 2 | 0.017 | 0.000 | 0.983 | 0.000 |
| 18 | 0 | 0.000 | 0 | 0.000 | 0.000 | 1.000 | Inf. |
| 19 | 0 | 0.000 | 2 | 0.017 | 0.000 | 0.982 | 0.000 |
| <u>Extreme Cut Points</u> |  |  |  |  |  |  |  |
| ≤6 | 10 | 0.086 | 6 | 0.051 | 0.085 | 0.949 | 1.667 |
| **7-8** | **47** | **0.402** | **9** | **0.077** | **0.402** | **0.923** | **5.222** |
| ≥9 | 60 | 0.513 | 102 | 0.872 | 0.513 | 0.128 | 0.588 |

<sup>a</sup> Developmental Language Disorder.
<sup>b</sup> Typically Developing.
\*The cutoffs for the best LH+ ratio for the measure are highlighted in bold. All scores with decimal values were rounded down to the nearest integer and were part of the corresponding range.

**Table 9.**
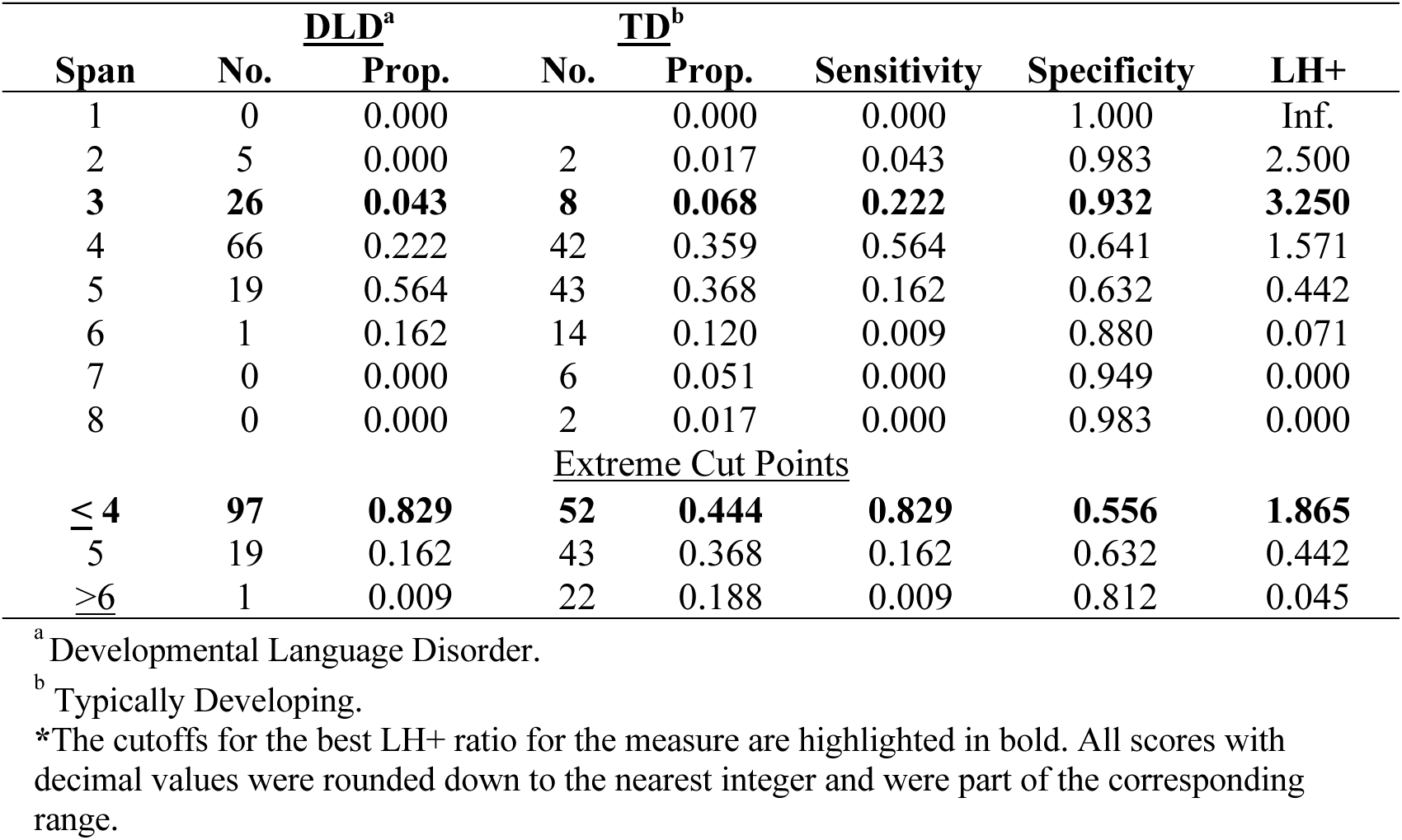
Likelihood (LH) ratio analysis for Digit Span for the 117/117 Cohort.

**Table 10.** Likelihood (LH) ratio analysis for RAN Mean RT for the 117/117 Cohort.

| RT msec | <u>DLD<sup>a</sup></u> |  | <u>TD<sup>b</sup></u> |  | Sensitivity | Specificity | LH+ |
| --- | --- | --- | --- | --- | --- | --- | --- |
|  | No. | Prop. | No. | Prop. |  |  |  |
| 500 | 0 | 0.000 | 1 | 0.009 | 0.000 | 0.991 | 0.000 |
| 600 | 2 | 0.017 | 7 | 0.060 | 0.017 | 0.940 | 0.286 |
| 700 | 8 | 0.068 | 27 | 0.231 | 0.068 | 0.769 | 0.296 |
| 800 | 23 | 0.197 | 37 | 0.316 | 0.197 | 0.684 | 0.622 |
| 900 | 26 | 0.222 | 27 | 0.231 | 0.222 | 0.769 | 0.963 |
| 1000 | 16 | 0.137 | 8 | 0.068 | 0.137 | 0.932 | 2.000 |
| 1100 | 10 | 0.086 | 3 | 0.026 | 0.085 | 0.974 | 3.333 |
| 1200 | 14 | 0.120 | 4 | 0.034 | 0.120 | 0.966 | 3.500 |
| 1300 | 3 | 0.026 | 2 | 0.017 | 0.026 | 0.983 | 1.500 |
| **1400** | **5** | **0.043** | **1** | **0.009** | **0.043** | **0.991** | **5.000** |
| 1500 | 3 | 0.026 | 0 | 0.000 | 0.026 | 1.000 | Inf. |
| 1600 | 2 | 0.017 | 0 | 0.000 | 0.017 | 1.000 | 0.000 |
| 1700 | 0 | 0.000 | 0 | 0.000 | 0.000 | 1.000 | Inf. |
| 1800 | 1 | 0.009 | 0 | 0.000 | 0.009 | 1.000 | Inf. |
| 1900 | 2 | 0.017 | 0 | 0.000 | 0.017 | 1.000 | Inf. |
| 2000 | 1 | 0.009 | 0 | 0.000 | 0.009 | 1.000 | Inf. |
| 2100 | 0 | 0.000 | 0 | 0.000 | 0.000 | 1.000 | Inf. |
| 2200 | 0 | 0.000 | 0 | 0.000 | 0.000 | 1.000 | Inf. |
| 2300 | 0 | 0.000 | 0 | 0.000 | 0.000 | 1.000 | Inf. |
| 2400 | 1 | 0.009 | 0 | 0.000 | 0.009 | 1.000 | Inf. |
| <u>Extreme Cut Points</u> |  |  |  |  |  |  |  |
| <1000 | 59 | 0.504 | 99 | 0.8462 | 0.50427 | 0.15385 | 0.59596 |
| **>1100** | **58** | **0.359** | **18** | **0.1538** | **0.49573** | **0.84615** | **3.22222** |

<sup>a</sup> Developmental Language Disorder.<sup>b</sup> Typically Developing.
\*The cutoffs for the best LH+ ratio for the measure are highlighted in bold. All scores with decimal values were rounded down to the nearest integer and were part of the corresponding range.

Haynes et al. (2006) argue that a measure must have an LHs value or above 20 to be used alone diagnostically as a conclusive test to rule in a disorder. For measures having LHs values below this range, Haynes et al. (2006) argue that while the measure is still valid, it must be used in conjunction with other diagnostic assessment tools for a valid diagnosis. These results show that none of the nine linguistic and/or cognitive processing measures had a sufficiently high LH value to be used alone in the diagnosis of DLD, although they all would be when used in conjunction with other measures to diagnose children with DLD.

Because the number of children with DLD identified at the highest LH+ differed across the nine measures, we asked if the *same* children with DLD were identified for the nine measures. We used twenty children identified by Digit Span Trials Correct because it had the highest LH+ and then asked how many of these children with DLD were also the *same* DLD children at the highest LH+ cut points for the remaining eight measures. For linguistic measures, there was little overlap. For Total Sentence Accuracy, only 2:6 children with DLD (LH+ = 6) were also from the group of twenty children identified by Digit Spans Trials Correct. For Noncanonical Sentence Accuracy, only 8:36 children with DLD (LH+ = 7.2) were the same. For Passive Sentence Accuracy, only 7:29 children with DLD (LH+ = 7.2) were also from the group of twenty. Similarly, for the working memory measures, for NRT, only 4:14 children with DLD (LH+ = 10) were from the group of twenty with DLD. For Working Memory Trials Correct, only 2:19 children with DLD (LH+ = 4.7) were the same. For the Working Memory Span task, only 3:10 children with DLD (LH+ = 5.0) were from the twenty with DLD. For Digit Span, 17:26 children with DLD (LH+ = 3.2) were the same. Finally, for RAN Mean RT, there was *no overlap* between the children identified as DLD on this measure (LH+ = 5.0) and the twenty children identified as DLD based on Digit Span Trials correct.

These results show that individually the LHs for each of the well established theoretically derived measures were moderately high, there was little consistency in which children were identified as DLD, however different children were identified as DLD across the measures. Specifically, the individual measures did not identify the *same* group of children as having DLD. In contrast, the elastic net regression model not only identified the subset of 71 features that were unique to a DLD-deficit profile but also obtained AUROCs of 87% - 88% in differentiating the 117 children with DLD from the 117 TD participants.

### Predicted probability score of DLD from Elastic Net Regression Models

One question is, was our elastic net model any more consistent in classifying the *same* individual child (e.g., internally consistent) with DLD over the 200 iterations of the model as compared to the LH analysis? To answer this, we examined the model-derived probability scores across 200 for the DLD and TD participants to determine if the *same* children received the same probability scores over the 200 model runs. The average of the model’s predictive probability score of a participant being DLD is shown in Figure 3A. The median and SD across the 200 predictive probability scores for each participant are shown in Figure 3B. The average predicted probabilities for DLD participants across the 200 iterations of the model for each participant were strongly correlated with the composite language z-score (*r* = .75, *p* < 0.001).

**Figure 3.**
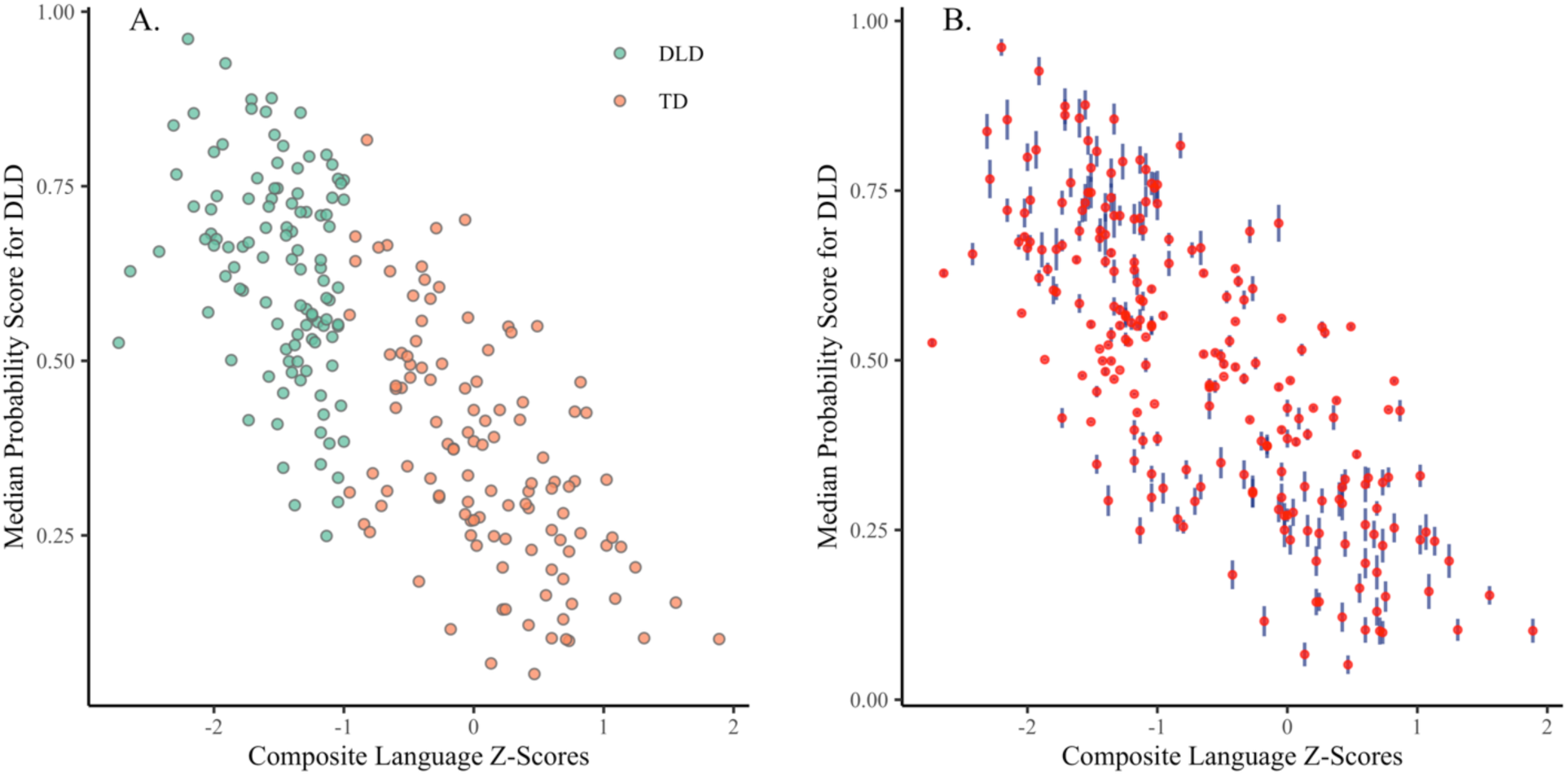
(A) Participants’ Median Predicted Probabilities of DLD Presence and their Composite Language Z-Scores. Each point represents a participant’s average probability score for having DLD, derived from 200 runs of elastic net logistic regression, with DLD (green) or TD (orange). The elastic net DLD-specific model assigned generally higher probabilities of DLD presence to participants with DLD and lower probabilities to participants with typical language throughout the runs, with some overlap in the .45- .55 probability range. (B) Participants’ Median Probabilities of DLD Presence and their Composite Language Z-Scores, along with the variability. The error line bar (blue line) is proportional to the standard deviation in probability scores across iterations, with a longer line reflecting greater variability. The variability across model iterations was low, with most minimal differences for TD and DLD participants, who received probability scores around .5, and slightly higher for the rest of both groups.

As can be seen in Figures 3A and B, the majority of the participants with DLD were assigned probability scores > .5, whereas most of the TD participants were assigned probability scores < .5, indicating that the elastic net models accurately separated the majority of the DLD and TD participants. There was however a subgroup of thirty-seven participants where the models consistently assigned probability scores of .45 - .55. This indicates that the models were uncertain as to group membership for these thirty-seven participants. Notably, the standard deviation of the probability scores for these participants was markedly lower than the remaining cohort (see Figure 3B; note that overall SD ranged from 0.0003 to 0.03). These thirty-seven participants (DLD = 18; TD = 19) were predominantly male (73%) as compared to the remaining sample (68% = male) and differed in age (p < .02) with the TD participants (7;0 – 11;25) being significantly younger than the DLD participants (9;10 – 11;5). The composite z-scores for these DLD and TD participants also differed significantly (p < .001), with the language composite z-score for these TD participants being significantly higher than for the DLD. The composite z-scores for the 18 DLD participants (-2.73 to -1.09) did not differ significantly for the other DLD participants (-2.46 to -1.00), (p = .41), The composite z-scores for the 19 TD participants (-.64 to .82) were however significantly lower than for the other TD participants (-.96 to 1.89), (p = .02), indicating that the language abilities of these 19 TD participants was lower than the remaining TD. Notably, the performance of the thirty-seven DLD and TD participants did not differ statistically from each other on the 9 experimental measures.

## Discussion

Our previous computational modeling work identified a unique DLD-specific deficit profile consisting of 9 linguistic and cognitive features (Sharma et al., under review; Sharma et al., in press). In this study, we examined the discriminative ability of each of the nine measures to rule in DLD in participants using likelihood ratios. Each measure showed intermediate-high separability, where five of the 9 measures yielded highest LH+ between 5+ and 10, and the remaining four yielded highest LH+ between 3 and 5. Since these measures did not reach the LH ratio >20, which is needed to confidently rule in or rule out DLD, the present analysis showed that these measures alone would be insufficient to differentiate DLD cases from TD. Similarly, of the 117 children with DLD, each measure’s LH identified no overlapping and non-consistent subsets of these children as having DLD.

The results from this study also show that the probability scores our elastic net model assigned to participants may have also identified a group of younger male participants with mild DLD, but who were not identified based on the polythetic classification approach used by Montgomery and colleagues. This is consistent with Fan et al.,’s (2017) findings. In addition to identifying a Williams Syndrome (WS)-specific neuroanatomical profile from a large number of MRI measures, where the predictive performance of their model accurately identified all of the 22 WS cohort, their model also identified undiagnosed “atypical” normal controls whose behavioral profiles were within normal limits but had partial deletions on the chromosome 7q11.23 as WS. Finally, this suggests that polythetic classification, originally used to identify children with DLD, wasn’t entirely sufficient or reliable either.

Taken together, the results of this study show that neither polythetic approach used by Montgomery and colleagues nor monothetic diagnostic approaches may be feasible for accurately diagnosing children with multidimensional disorders such as DLD. Specifically, our results show that computational approaches, such as elastic net models, can capture the small and often non-significant contributions of the disorder’s multiple features. This suggests that viewing DLD as a multidimensional structure may be a more viable approach since it is not a single feature emerging from a single cause; rather, subtle differences and deficits across multiple domains, such as verbal working memory, language comprehension, and processing speed, accumulate in a modest yet meaningful manner to give rise to heterogeneous, complex DLD disorders.

The findings of this study align well with recent work by Bishop and McDonald (2009), who showed that while language test measures provide valuable information for identifying children in need, incorporating multiple aspects, such as parental and teacher reports, enhanced diagnostic accuracy. Her later study, which included a large panel of DLD researchers from various English-speaking countries and fields of practice to reach consensus on terminology (Bishop, 2017), also highlighted that neither a single cognitive challenge—within working memory, nonverbal intelligence, or motor coordination —nor a single language difference—within morphology, syntax, semantics, or pragmatics—is uniformly present in participants with DLD. Overall, the consensus was that no single measure provides a robust psychometric foundation, but a multitude of assessment measures should be used to identify the disorder.

A significant limitation of this study is that it relies on the secondary dataset and diagnostic classification criteria. As such, it inherits the theoretical constraints from the original Montgomery et al project. First, although the database included a high-density sample of 71 language and cognition measures, including reaction time, accuracy, and error patterns for working memory, canonical and non-canonical sentence comprehension, attentional control, etc., from school-aged children, it lacked morphosyntactic measures. Second, while we showed that the repeated elastic net logistic regression was robust in predicting the likelihood of DLD presence in participants from the sample on average, to date, the generalizability of the model has not been examined an independent dataset and separate clinical sample.

## Author Contributions

Conceptualization, J.L.E. and S.S.; methodology, J.L.E., R.B.G., J.W.M, R.G., and S.S.; software, S.S.; validation, J.L.E., S.S., and R.G.; formal analysis, J.L.E. and S.S.; investigation, J.L.E., S.S., R.G., R.B.G., and J.W.M; resources, J.L.E.; data curation, J.L.E., R.B.G., and J.W.M; writing—original draft preparation, J.L.E. and S.S.; writing—review and editing, J.L.E. and S.S.; visualization, J.L.E. and S.S.; supervision, J.L.E.; project administration, J.L.E. and R.G..; funding acquisition, J.L.E., J.W.M, and R.B.G. All authors have read and agreed to the published version of the manuscript.

## Institutional Review Board Statement

The study was conducted in accordance with the Declaration of Helsinki as well as the guidelines of the University Institutional Review Board (IRB), which approved the protocol (IRB-24-258).

## Patient Informed Consent Statement

Informed consent was obtained from all subjects involved in the study.

## Data Availability Statement

The data that support the findings of this study are available from the corresponding author upon reasonable request.

## Acknowledgments

This research was conducted using the secondary database collected by three investigators: James Montgomery, Julia Evans, and Ronald Gillam. We are grateful for their foundational contributions. All authors have reviewed and edited the output and take full responsibility for the content of this publication.

## References

Ahufinger, N. et al. (2021) “Consistency of a Nonword Repetition Task to Discriminate Children with and without Developmental Language Disorder in Catalan-Spanish and European Portuguese Speaking Children,” Children (Basel, Switzerland), 8(2), p. 85. Available at: 10.3390/children8020085.

American National Standards Institute. (1997). Specifications of audiometers (ANSI/ANS 8.3-1997, R2003). Author.

Bishop, D.V.M. (2017) “Why is it so hard to reach agreement on terminology? The case of developmental language disorder (DLD),” International Journal of Language & Communication Disorders, 52(6), pp. 671–680. Available at: 10.1111/1460-6984.12335.

Bishop, D.V.M. and McDonald, D. (2009) “Identifying language impairment in children: combining language test scores with parental report,” International Journal of Language & Communication Disorders, 44(5), pp. 600–615. Available at: 10.1080/13682820802259662.

Botting, N. et al. (2016) “Emotional health, support, and self-efficacy in young adults with a history of language impairment,” The British Journal of Developmental Psychology, 34(4), pp. 538–554. Available at: 10.1111/bjdp.12148.

Conti-Ramsden, G. et al. (2013) “Adolescents with a history of specific language impairment (SLI): strengths and difficulties in social, emotional and behavioral functioning,” Research in Developmental Disabilities, 34(11), pp. 4161–4169. Available at: 10.1016/j.ridd.2013.08.043.

Dollaghan, C. and Campbell, T.F. (1998) “Nonword repetition and child language impairment,” Journal of speech, language, and hearing research: JSLHR, 41(5), pp. 1136–1146. Available at: 10.1044/jslhr.4105.1136.

Ellis Weismer, S., et al. (2000) “Nonword repetition performance in school-age children with and without language impairment,” Journal of speech, language, and hearing research: JSLHR, 43(4), pp. 865–878. Available at: 10.1044/jslhr.4304.865.

Fan, C.C., Brown, T.T., Bartsch, H., Kuperman, J.M., Hagler Jr, D.J., Schork, A., Searcy, Y., Bellugi, U., Halgren, E. and Dale, A.M., 2017. Williams syndrome-specific neuroanatomical profile and its associations with behavioral features. NeuroImage: Clinical, 15, pp.343–347.

Florkowski, C.M. (2008) Sensitivity, Specificity, Receiver-Operating Characteristic (ROC) Curves and Likelihood Ratios Communicating the Performance of Diagnostic Tests. The Clinical Biochemist Reviews, 29, S83–S87. *- References - Scientific Research Publishing* (no date). Available at: https://www.scirp.org/reference/referencespapers?referenceid=2386901 (Accessed: January 17, 2026).

Friedman, J. et al. (2008) “glmnet: Lasso and Elastic-Net Regularized Generalized Linear Models.” Available at: 10.32614/CRAN.package.glmnet.

Friedman, J., Hastie, T. and Tibshirani, R. (2010) “Regularization Paths for Generalized Linear Models via Coordinate Descent,” Journal of Statistical Software, 33(1), pp. 1–22.

Hubert-Dibon, G. et al. (2016) “Health-Related Quality of Life for Children and Adolescents with Specific Language Impairment: A Cohort Study by a Learning Disabilities Reference Center,” PloS One, 11(11), p. e0166541. Available at: 10.1371/journal.pone.0166541.

Girbau, D. and Schwartz, R.G. (2007) “Non-word repetition in Spanish-speaking children with Specific Language Impairment (SLI),” International Journal of Language & Communication Disorders, 42(1), pp. 59–75. Available at: 10.1080/13682820600783210.

Hammill DD, Newcomer PL. (2008) Test of Language Development-Intermediate: Fourth Edition. Austin, TX: Pro-Ed.

Haynes, R.B., Sackett, D.L., Guyatt, G.H. and Tugwell, P., 2006. How to do clinical practice research: a new book and a new series in the Journal of Clinical Epidemiology. Journal of clinical epidemiology, 59(9), pp.873–875.

Leonard, L. (2014). Children with specific language impairment. Cambridge, MA: MIT Press.

Marinis, T. and van der Lely, H.K.J. (2007) “On-line processing of wh-questions in children with G-SLI and typically developing children,” International Journal of Language & Communication Disorders, 42(5), pp. 557–582. Available at: 10.1080/13682820601058190.

Montgomery, J.W. et al. (2016) “‘Whatdunit?’ Developmental changes in children’s syntactically based sentence interpretation abilities and sensitivity to word order,” Applied Psycholinguistics, 37(6), pp. 1281–1309. Available at: 10.1017/S0142716415000570.

Montgomery, J.W. et al. (2018) “Structural Relationship Between Cognitive Processing and Syntactic Sentence Comprehension in Children With and Without Developmental Language Disorder,” Journal of Speech, Language, and Hearing Research : JSLHR, 61(12), pp. 2950–2976. Available at: 10.1044/2018_JSLHR-L-17-0421.

Montgomery, J.W. et al. (2017) “‘Whatdunit?’ Sentence Comprehension Abilities of Children With SLI: Sensitivity to Word Order in Canonical and Noncanonical Structures,” Journal of Speech, Language, and Hearing Research, 60(9), pp. 2603–2618. Available at: 10.1044/2017_JSLHR-L-17-0025.

Rice, M.L., Earnest, K.K. and Hoffman, L. (2023) “Longitudinal Grammaticality Judgments of Tense Marking in Complex Questions in Children With and Without Specific Language Impairment, Ages 5-18 Years,” Journal of speech, language, and hearing research: JSLHR, 66(10), pp. 3882–3906. Available at: 10.1044/2023_JSLHR-22-00507.

Roid, G.H. and Miller, L.J., 1997. Leiter international performance scale-revised: Examiners manual (Vol. 10). Wood Dale, IL: Stoelting.

Ruscio, J., Ruscio, A.M. and Carney, L.M. (2011) “Performing Taxometric Analysis to Distinguish Categorical and Dimensional Variables,” Journal of Experimental Psychopathology, 2(2), pp. 170–196. Available at: 10.5127/jep.010910.

Sackett, D. L. (1991) Clinical epidemiology : a basic science for clinical medicine. 2nd ed. Boston: Little, Brown.

Semel, E.M., Wiig, E.H. and Secord, W., 2003. Clinical evaluation of language fundamentals.

Sharma, S., Golden, R. M., Montgomery, J., Gillam, R. B., & Evans, J. L. (in press). Using Predictive Performance from an Elastic Net Regression to Classify Developmental Language Disorder (DLD). Annals of Otolaryngology and Rhinology

Sharma, S., Golden, R. M., Montgomery, J., Gillam, R. B., & Evans, J. L. (2025). Developmental Language Disorder (DLD): Specific Deficit Profile and its Associated Language and Cognitive Processing Features [Manuscript submitted for publication]. Department of Speech, Language, and Hearing Sciences, The University of Texas at Dallas

Tomblin, J.B., Records, N.L. and Zhang, X. (1996) “A system for the diagnosis of specific language impairment in kindergarten children,” Journal of Speech and Hearing Research, 39(6), pp. 1284–1294. Available at: 10.1044/jshr.3906.1284.

Tucker, L.R. and Lewis, C. (1973) “A reliability coefficient for maximum likelihood factor analysis,” Psychometrika, 38(1), pp. 1–10. Available at: 10.1007/BF02291170.

Wallace, G. and Hammill, D.D., 2002. Comprehensive receptive and expressive vocabulary test. Austin, TX: Pro-Ed.

Woodcock, R., McGrew, K., & Mather, N. (2001). Woodcock- Johnson III Test of Cognitive Abilities. Itasca, IL: Riverside Publishing.

